# Ethnic Disparities in Hospitalization for COVID-19: a Community-Based Cohort Study in the UK

**DOI:** 10.1101/2020.05.19.20106344

**Authors:** Camille Lassale, Bamba Gaye, Mark Hamer, Catharine R. Gale, G. David Batty

## Abstract

**Importance:** Differentials in COVID-19 incidence, hospitalization and mortality according to ethnicity are being reported but their origin is uncertain.

**Objective:** We aimed to explain any ethnic differentials in COVID-19 hospitalization based on socioeconomic, lifestyle, mental and physical health factors.

**Design:** Prospective cohort study with national registry linkage to hospitalisation for COVID-19.

**Setting:** Community-dwelling.

**Participants:** 340,966 men and women (mean age 56.2 (SD=8.1) years; 54.3% women) residing in England from the UK Biobank study.

**Exposures:** Ethnicity classified as White, Black, Asian, and Others.

**Main Outcome(s) and Measure(s):** Cases of COVID-19 serious enough to warrant a hospital admission in England from 16-March-2020 to 26-April-2020.

**Results:** There were 640 COVID-19 cases (571/324,306 White, 31/4,485 Black, 21/5,732 Asian, 17/5,803 Other). Compared to the White study members and after adjusting for age and sex, Black individuals had over a 4-fold increased risk of being hospitalised (odds ratio; 95% confidence interval: =4.32; 3.00-6.23), and there was a doubling of risk in the Asian group (2.12; 1.37, 3.28) and the ‘other’ non-white group (1.84; 1.13, 2.99). After controlling for 15 confounding factors which included neighbourhood deprivation, education, number in household, smoking, markers of body size, inflammation, and glycated haemoglobin, these effect estimates were attenuated by 33% for Blacks, 52% for Asians and 43% for Other, but remained raised for Blacks (2.66; 1.82, 3.91), Asian (1.43; 0.91, 2.26) and other non-white groups (1.41; 0.87, 2.31).

**Conclusions and Relevance:** Our findings show clear ethnic differences in risk of hospitalization for COVID-19 which do not appear to be fully explained by known explanatory factors. If replicated, our results have implications for health policy, including the targeting of prevention advice and vaccination coverage.

**Key points:** *Question:* What explains ethnic differences in rates of hospitalisation for COVID-19?

*Finding:* In a large, community-based cohort, Black and Asian individuals had a markedly higher risk of hospitalisation. After adjustment for socioeconomic, lifestyle, comorbidities, and biomarkers, Black individuals still experienced more than a doubling of risk compared to white individuals though the effect for the Asian group was diminished.

*Meaning:* In England, the marked ethnic disparities in the risk of hospitalisation for COVID-19, if replicated, has implications for health policy, including the targeting of prevention advice and vaccination coverage.

## Introduction

Ethnic disparities in health have traditionally been examined for non-communicable disease, chiefly cardiovascular disease, however, there is emerging evidence that COVID-19 might disproportionately affect people from ethnic backgrounds.^1,2^ In the UK, inequalities in COVID-19 in prognostic studies have been reported such that, in cohorts of hospitalised patients, minority groups appear to have the greatest risk of progression to intensive care and death.^3^ In the US, a pooling of hospital data from 38 states also shows that minorities have a greater rate of deaths involving COVID-19, and this is particularly so for African-Americans.^4,5^

With neighbourhood deprivation and comorbidity only partially explain these ethnic differentials,^3^ other causes need to be examined. These include individual socioeconomic disadvantage such as education, overrepresentation of minorities in in public-facing occupations, overcrowded living and working conditions, and greater prevalence of unhealthy lifestyle and chronic disease.^1,6-8^ Mental health problems, also more common in minorities,^9,10^ may also be related to infection and severity of respiratory infections via impaired innate and adaptive immunity.^11,12^ Finally, biological differences, such as impaired immunologic response functioning,^13^ are amplified in the present of racism and chronic stress.

With existing studies focusing on disease prognosis, it is unclear if people from ethnic groups also experience an elevated risk of disease onset, and, if so, what explains this burden. Accordingly, our aim was to assess the ethnic differences in serious cases of COVID-19 in a well-characterized, large, community-based cohort study in the UK, and investigate which underlying factors drive the observed associations.

## Methods

### Study Population

We used data from UK Biobank, a prospective cohort study, the sampling and procedures of which have been well described.^14^ Baseline data collection took place between 2006 and 2010 across twenty-two research assessment centres in the UK giving rise to a sample of 502,655 people aged 40 to 69 years (response rate 5.5%).^14^ Ethical approval was received from the North-West Multi-centre Research Ethics Committee, and the research was carried out in accordance with the Declaration of Helsinki of the World Medical Association, and participants gave informed consent. For the present analysis, participants residing in Scotland, Wales, or Northern Ireland were excluded as COVID-19 test data were only available for England.

### Hospitalisation for COVID-19

Provided by Public Health England, data on COVID-19 status covered the period from 16^th^ March to 26^th^April 2020 (http://biobank.ndph.ox.ac.uk/showcase/field.cgi?id=40100), during which testing was largely restricted to those with symptoms in hospital. COVID-19 tests were performed on samples from combined nose/throat swabs using real time polymerase chain reaction (RT-PCR) in accredited laboratories.^15^ These data can therefore be regarded as a proxy for hospitalisations for severe COVID-19 cases for England only.

### Ethnicity

Ethnicity was self-reported at baseline assessment and included 6 categories: White (including White British, White Irish, any other white background), Mixed (White and Black Caribbean, White and Black African, White and Asian, any other mixed background), Asian or Asian British (thereafter termed “Asian”, including Indian, Pakistani, Bangladeshi, any other Indian background), Black or Black British (“Black”, Caribbean, African, any other Black background), Chinese, and Other. To maintain statistical power in our analyses, we grouped together Chinese, Mixed and Other under the “Other” category.

#### Covariates

All variables were obtained at baseline and were grouped into 4 clusters.

#### Socioeconomic factors

Socioeconomic factors included highest educational attainment, household income, occupation, number of people living in the household, and the Townsend index of area deprivation^16^ (higher values denote deprivation). We created binary variables for education (university degree yes/no), total household income before tax (<18,000 GBP or above), occupation (non-manual, manual). Size of the household had four groups (living alone; with two people; with three people; and four or more).

#### Lifestyle Measures

Physical activity, smoking, and alcohol consumption were assessed by questionnaire. Participants were categorised into never, previous, and current smokers. We grouped alcohol intake into three categories: never/rarely, and below or above current UK guidelines (≥14 units in women and ≥21 units in men). Leisure time physical activity was assessed using the short form version of the International Physical Activity Questionnaire (IPAQ). ^17^ Measuring duration and frequency of moderate-to-vigorous physical activity in the last week, data were grouped in 3 categories: inactive, somewhat active below the guidelines, and meeting activity guidelines (≥150 min/week moderate-to-vigorous physical activity or ≥75 min/week vigorous activity).^18^

#### Comorbidities

Body weight was measured using Tanita BC418MA scales and standing height using a Seca height measure, and body mass index (BMI) calculated [weight (kilograms)/height^2^ (meters^2^) squared]. Waist and hip circumference were measured with a non-elastic tape, and their ratio computed. The following self-reported physician diagnosed chronic diseases were used: cardiovascular diseases (heart attack, angina, stroke), chronic bronchitis and diabetes. Hypertension was defined as elevated measured blood pressure (≥140/90 mmHg) and /or use of anti-hypertensive medication. We used two indicators of mental health: contact with a psychiatrist for any disorder and symptoms of psychological distress as measured using the four-item version of the Patient Health Questionnaire (PHQ-4) in which scores ranged from 0-12 (categorised as 0, 1-2, ≥3 [high]). A verbal numerical reasoning task was used as a marker of cognitive function.^12^

#### Biomarkers

Non-fasting venous blood samples were drawn and assayed for C-reactive protein, glycated haemoglobin, and total and high-density lipoprotein cholesterol.^19,20^ Forced expiratory volume in 1 second, a marker of lung function, was quantified using spirometry with the best of three technically satisfactory exhalations used.

## Statistical Analyses

We fitted logistic regression models to estimate odds ratios and 95% confidence intervals for associations between ethnicity and hospitalization for COVID-19. With the outcome being rare, odds ratios (OR) can be interpreted as relative risks (RR). To assess the contribution of factors to the ethnic differences, we used a simple coefficient attenuation approach. Beginning with a comparator model where ORs were adjusted for age and sex, we subsequently fitted 5 models corresponding to groups of covariates: 1) socioeconomic, 2) lifestyle, 3) comorbidities, 4) biomarkers, and 5) all covariates. Attenuation expressed in percentage was calculated as 100*(β_model x_ – β_base model_)/ β_base model_. Because the aim is to compare attenuation of ORs by inclusion of various sets of factors, we selected all participants with non-missing values to run all five models. In a first sensitivity analysis, we present the estimates in samples with the maximum number of observations for each model. The cognitive function variable was only available in a subset of participants, therefore we present as a sensitivity analysis for the complete-case model with and without this factor. We also conducted the analysis separately for men and women. Finally, we also present results where covariates were imputed using multiple imputations by chain equations with 2 datasets.

## Results

Ethnicity data were available for 428,494 participants (235,528 women, 55%) who were alive prior to COVID-19 testing (up to 5 March 2020). The main analytical sample comprised 340,966 participants (640 COVID-19 cases) with complete data on the core set of covariates listed in Table 1. After adjusting for age and sex, compared to White participants, being from a Black ethnic background was associated with over a four-fold risk of hospitalization for COVID-19 (odds ratio; 95% confidence interval: 4.32; 3.00-6.23), while a doubling was apparent in Asian (2.12; 1.37, 3.28) and Other (1.84; 1.13, 2.99) (Table 1). Gradual attenuation of the association after inclusion of groups of confounders can be seen in Figure 1. The greatest attenuations were observed when socioeconomic factors were added to the multivariable model: 24.5% for Blacks, 31.9.3% for Asians, and 30.0 % for Others. After further control for lifestyle factors, co-morbidities, and biomarkers, relationships were attenuated by 33.0% for Blacks, 52.2% for Asians and 43.0% for Others compared to the base model. There was, however, still evidence of associations, most obviously for Blacks (2.66; 1.82, 3.91).

**Table 1.** Baseline characteristics of participants across ethnic groups, UK Biobank.

|  | White | Asian | Black | Other |
| --- | --- | --- | --- | --- |
| N | 403196 | 9260 | 7734 | 8304 |
| COVID-19 cases (n (%)) | 571 (0.19) | 21 (0.50) | 31 (0.70) | 17 (0.34) |
| Women (n (%)) | 221,656 (55.0) | 4,350 (47.0) | 4,507 (58.3) | 5,015 (60.4) |
| Age, years (mean (SD)) | 56.6 (8.0) | 53.2 (8.4) | 51.8 (8) | 52.1 (7.9) |
| *Percent* |  |  |  |  |
| Higher education | 32.1 | 41 | 33.9 | 45.8 |
| Household $\geq 4$ people | 18.1 | 51.7 | 31.9 | 32.2 |
| Neighborhood deprivation Highest quintile | 17.9 | 37.8 | 63.7 | 44.2 |
| Physical inactivity | 18 | 23.9 | 20.4 | 20.8 |
| Alcohol intake |  |  |  |  |
| Within guideline | 36.8 | 19.5 | 24.9 | 24.3 |
| Never/rarely | 29.2 | 72.1 | 65.1 | 61.2 |
| Heavy drinking | 34 | 8.3 | 10 | 14.5 |
| Smoking |  |  |  |  |
| Never | 54.5 | 77.6 | 70.7 | 60.4 |
| Past smoker | 35.6 | 13 | 17.3 | 25.7 |
| Current smoker | 9.9 | 9.4 | 12 | 13.8 |
| Hypertension | 58.2 | 56.4 | 62.4 | 49.3 |
| Diabetes | 4.5 | 16.8 | 11.1 | 7.9 |
| Cardiovascular disease | 5.3 | 7.8 | 4.6 | 4 |
| Chronic bronchitis | 1.4 | 0.9 | 0.6 | 0.9 |
| Seen a psychiatrist | 11.4 | 10.2 | 8.6 | 12.6 |
| Psychological distress (PHQ4$\geq 3$) | 22.9 | 41.7 | 36.5 | 36.2 |
| *Mean (SD)* |  |  |  |  |
| BMI, kg/m<sup>2</sup> | 27.4 (4.7) | 27.2 (4.4) | 29.5 (5.4) | 27.0 (5.0) |
| Waist to hip ratio | 0.87 (0.09) | 0.9 (0.08) | 0.87 (0.08) | 0.87 (0.08) |
| C-reactive protein (mg/L) | 2.50 (4.18) | 2.79 (3.99) | 2.78 (4.4) | 2.37 (3.99) |
| HbA1c (mmol/mol) | 35.8 (6.3) | 40.5 (10.3) | 39.3 (10.0) | 37.5 (8.2) |
| Cholesterol (mmol/L) | 5.72 (1.14) | 5.33 (1.12) | 5.25 (1.09) | 5.53 (1.11) |
| HDL-cholesterol (mmol/L) | 1.46 (0.38) | 1.26 (0.32) | 1.44 (0.36) | 1.42 (0.38) |
| Forced expiratory volume in 1 sec (L) | 2.85 (0.79) | 2.23 (0.73) | 2.33 (0.73) | 2.54 (0.76) |

**Figure 1.**
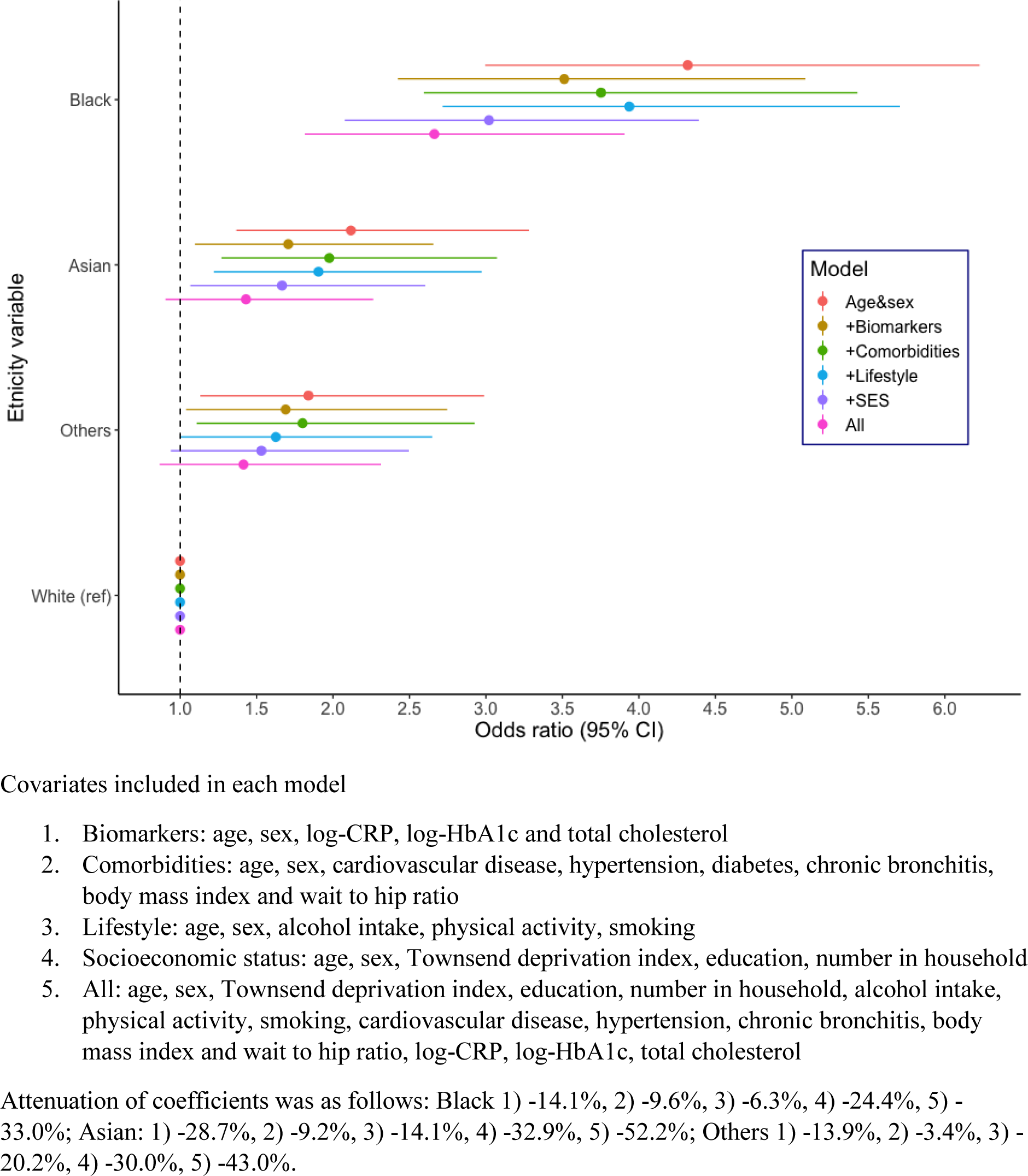
Association between ethnicity and hospitalization for COVID-19 in UK Biobank, n=324, 877.

Effects for Asians (1.43; 0.91, 2.26) and Others (1.41; 0.87, 2.31), while raised, were not statistically significant at conventional levels (Table 2). In sex-specific analysis (Supplemental Table 1), we found that ORs for Black men (multivariable OR compared to white men: 3.51; 2.11, 5.81) were greater than for Black women (1.93; 1.07, 3.48, compared to white women). Contrarily, ORs for people from an Asian background were lower and weakened to a greater extent after inclusion of the full set of covariates for men than for women (attenuation by 72% for men, multivariable OR: 1.16; 0.60, 2.32, attenuation by 38% in women, OR 1.91; 1.01, 3.62).

**Table 2.** Multiply-adjusted odds ratios and 95% confidence intervals for the relation of baseline characteristics with hospitalization for COVID-19.

|  | Age- and sex-adjusted |  |  | Multiply-adjusted <sup>a</sup> |  |  |  |
| --- | --- | --- | --- | --- | --- | --- | --- |
| N cases / N total<br>640 / 340,966 | OR | 95% CI | p-value | OR | 95% CI | p-value | Attenuation<br>% <sup>c</sup> |
| Ethnicity (reference=White) |  |  |  |  |  |  |  |
| Black (29 / 4,516) | 4.32 | (3.00 - 6.23) | <0.001 | 2.66 | (1.82 - 3.91) | <0.001 | -33.0 |
| Asian (21 / 5,753) | 2.12 | (1.37 - 3.28) | 0.001 | 1.43 | (0.91 - 2.26) | 0.125 | -52.2 |
| Other (17 / 5,820) | 1.84 | (1.13 - 2.99) | 0.014 | 1.42 | (0.87 - 2.31) | 0.166 | -43.0 |
| Age (years) | 1.02 | (1.01 - 1.03) | 0.001 | 1.02 | (1.01 - 1.03) | 0.003 |  |
| Male (vs female) | 1.56 | (1.34 - 1.83) | <0.001 | 1.15 | (0.92 - 1.44) | 0.219 |  |
| Lower education |  |  |  | 1.15 | (0.96 - 1.37) | 0.131 |  |
| Number in household (ref=2) |  |  |  |  |  | 0.001<sup>b</sup> |  |
| One person |  |  |  | 1.15 | (0.93 - 1.43) | 0.195 |  |
| 3 people |  |  |  | 1.22 | (0.97 - 1.55) | 0.093 |  |
| 4 people or more |  |  |  | 1.59 | (1.26 - 2.01) | <0.001 |  |
| Townsend score (ref=least deprived, Q1) |  |  |  |  |  | <0.001<sup>b</sup> |  |
| Q2 |  |  |  | 1.00 | (0.76 - 1.33) | 0.989 |  |
| Q3 |  |  |  | 0.99 | (0.75 - 1.31) | 0.937 |  |
| Q4 |  |  |  | 1.24 | (0.95 - 1.62) | 0.116 |  |
| Q5 |  |  |  | 1.67 | (1.30 - 2.16) | <0.001 |  |
| Physical activity (ref=meeting guideline) |  |  |  |  |  | 0.045<sup>b</sup> |  |
| Active >10min not reaching guideline |  |  |  | 0.93 | (0.77 - 1.13) | 0.466 |  |
| Inactive |  |  |  | 1.22 | (1.00 - 1.48) | 0.049 |  |
| Alcohol (ref=within guideline) |  |  |  |  |  | 0.041<sup>b</sup> |  |
| Never/very rarely drink |  |  |  | 1.30 | (1.07 - 1.59) | 0.01 |  |
| Intake above guideline |  |  |  | 1.10 | (0.90 - 1.34) | 0.368 |  |
| Smoking (ref=never smoker) |  |  |  |  |  | 0.008<sup>b</sup> |  |
| Ex-smoker |  |  |  | 1.30 | (1.10 - 1.55) | 0.003 |  |
| Current smoker |  |  |  | 1.25 | (0.96 - 1.62) | 0.095 |  |
| Body mass index (kg/m<sup>2</sup>) |  |  |  | 1.03 | (1.02 - 1.05) | <0.001 |  |
| Waist-to-hip ratio (0.1 unit increase) |  |  |  | 1.25 | (1.09 - 1.42) | 0.001 |  |
| Hypertension |  |  |  | 0.98 | (0.82 - 1.17) | 0.84 |  |
| Cardiovascular disease |  |  |  | 1.06 | (0.79 - 1.42) | 0.705 |  |
| Chronic bronchitis |  |  |  | 1.34 | (0.81 - 2.21) | 0.259 |  |
| Ever seen a psychiatrist |  |  |  | 1.24 | (0.99 - 1.55) | 0.057 |  |
| log-CRP (1 unit increase) |  |  |  | 1.05 | (0.92 - 1.19) | 0.477 |  |
| log-HbA1c (1 unit increase) |  |  |  | 1.60 | (1.02 - 2.52) | 0.043 |  |
| Cholesterol (mmol/L) |  |  |  | 0.90 | (0.84 - 0.97) | 0.004 |  |

<sup>a</sup> Estimates are all mutually adjusted,
<sup>b</sup> p-trend,
<sup>c</sup> Attenuation from the age and sex adjusted estimate to the multivariable adjusted estimate

In the maximum sample approach, the same pattern was observed (Supplemental Table 2). In a reduced sample of 116,990 individuals with available cognitive test score, associations were further attenuated after inclusion of this variable in the model, which displayed a strong association with COVID-19 hospitalization (Supplemental Table 3). Finally, using multiple imputation, fully adjusted ORs were as follows: Black 2.53; 95% CI 1.87, 3.42; Asian 1.63; 1.17, 2.26; Others 1.44; 0.97, 2.12 (Supplemental Table 4).

## Discussion

In a large community-dwelling cohort of over 400,000 individuals, we found that ethnic minority groups in England experience a higher risk of COVID-19 hospitalization such that the greatest risk effect observed for people of Black ethnic origin. The observed associations were attenuated but remained strong after adjustment for socioeconomic, lifestyle and health-related factors.

This work complements emerging evidence from various countries, in particular the USA and the UK, in large ethnically diverse populations, of disproportionately high rates of death involving COVID-19 in ethnic minority groups.^2,8^ There are several hypotheses that might explain these disparities. Firstly, minority ethnic groups are more likely to be in public-facing, service-based occupations which may mean they are less able to take effective physical distancing measures. Secondly, they are more likely to be of low income, in precarious contracts or self-employed, and to be living in intergenerational crowded households.^2^ Moreover, if not legally resident, migrants may be fearful of accessing official health care services.^7^ In the present analysis, we observed that household composition and neighbourhood deprivation are strong predictors of COVID-19 hospitalization and partially attenuated the association between ethnicity and COVID-19.

It is also known that there are disparities in lifestyle and ill health, mental and physical, across ethnic groups,^21,22^ which may explain susceptibility to a severe COVID-19 infection. However, although being important predictors, lifestyle, morbidity, biomarkers and mental health again only partially diminished the association between the infection and ethnicity. Adding biomarkers into the model did have explanatory power, particularly in men, which is perhaps due to the high prevalence of diabetes and elevated HbA1c in the Asian population, and may also be due to presence of low grade inflammation as seen in the higher C-reactive protein levels. Another important result is the strength of the association between mental illness and COVID-19, and how taking into account cognitive function attenuated the association with all ethnic groups. However, markers of mental health, alongside inflammation, which may result from racism or other stressors experienced more often by ethnic minority, did not fully explain the association, although specific measures of chronic stress and discrimination would have been more accurate. Given the rise of evidence, calls have been made to investigate the role of highly prevalent chronic conditions, such as obesity and diabetes, in explaining the observed ethnic disparities in COVID-19.^23^ With these results, we provide new insights into this observed relationship.

This is the first study of disease onset and one which takes into account an extensive set of potential confounders and mediators, spanning individual and neighbourhood socioeconomic factors, lifestyle and markers of mental and physical health. The study has other strengths, including being based on a well-characterized large community-based sample. Additionally, study members were linked to objective measurement of the disease as opposed to self-report with confirmation of COVID-19 status being based on biological samples using PCR methodology, considered to be the gold standard. The study is not without its weaknesses. First, due to the absence of systematic testing across the UK, these data come from hospital records, therefore reflect only patients with a manifestation of the disease severe enough to require inpatient admission into hospital. Some cases of COVID-19 could also have been captured in patients originally hospitalized for reasons other than the infection. Second, the UK Biobank cohort is not representative of the general UK population. Therefore, absolute prevalence and risks should not be interpreted as such, but an aetiological investigation of risk factor association such as the present study are likely to be generalizable.^24^ However, it is important to keep in mind that double selection of the sample - UK Biobank participants are not representative from the general population, and we selected a non-missing analytical sample within the cohort - may lead to collider bias.^25^ This means that conditioning on factors associated with the selection of the sample can distort or induce spurious associations. For example, this is likely to have been the case in studies finding that current smokers appear protected against COVID-19.^26^ In the present study, smoking (in particular ex-smokers) was associated with greater risk of COVID-19 hospitalization, somewhat ruling out collider bias. Third, despite using an extensive set of socioeconomic factors, both at individual and area level, we failed to capture some features that may be particularly relevant to the ethnic differences observed in the COVID-19 pandemic context: occupation did not classify between public facing occupations, not only health professionals, but also supermarket clerks, bus drivers or couriers. The number of people in the household, while a proxy for overcrowding, does not capture intergenerational co-living. Finally, exposure data were collected a few years ago (2006-2010) and participants’ living circumstances may have changed. Also, we excluded study members who had died prior to 5^th^ March 2020 because they could not contribute to the risk set, however, ascertainment of COVID-19 hospitalisation did not reliably begin until 16^th^ March. It is unlikely, however, that the absence of vital status data for this 11-day period would have biased our effect estimates.

To conclude, in England, the observed ethnic disparities in hospitalization for COVID-19 was strong, in particular comparing Black and White individuals, and not fully explained by an extensive set of socioeconomic, lifestyle and health disparities. If replicated, this has implications for health policy, including the targeting of prevention advice and vaccination coverage. Further research is needed to better understand the underlying mechanisms driving the racial/ethnic disparities in hospitalization for COVID-19 observed in our study.

## Data Availability

Access to data: Data from UK Biobank (http://www.ukbiobank.ac.uk/) are available to bona fide researchers upon application. This research has been conducted using the UK Biobank Resource under Application 10279.

## Acknowledgements

Acknowledgement:

We thank the UK Biobank study members for their generosity in participating.

## Access to data

Data from UK Biobank (http://www.ukbiobank.ac.uk/) are available to bona fide researchers upon application. This research has been conducted using the UK Biobank Resource under Application 10279.

## Contributions

GDB and CL generated the idea for the present paper; CRG prepared the data set; CL carried out all the data analyses; CL, BG, and GDB wrote the manuscript; All authors commented on an earlier version of the manuscript. CL will act as guarantors for this work. The corresponding author attests that all listed authors meet authorship criteria and that no others meeting the criteria have been omitted.

